# Generational differences in physical health and disability in the United States and Europe

**DOI:** 10.1101/2024.01.16.24301347

**Authors:** Laura Gimeno, Alice Goisis, Jennifer B. Dowd, George B. Ploubidis

**Author notes:** Corresponding author. UCL Centre for Longitudinal Studies, 55-59 Gordon Square, London WC1H 0NU.

## Abstract

**Objectives:** Declines in mortality have typically been associated with improvements in physical health across generations. While life expectancy in most high-income countries continues to increase, there is evidence that younger generations, particularly in the United States (US), are less healthy than previous generations at the same age. We compared generational trends in physical health in the US, England, and continental Europe to explore whether other regions have experienced a similar pattern of worsening health across cohorts.

**Methods:** Using data from nationally representative studies of adults aged ≥50 years from the US (Health and Retirement Study, *n*=26,939), England (English Longitudinal Study of Ageing, *n*=14,992) and 11 continental European countries (Survey of Health, Ageing and Retirement in Europe, *n*=72,595), we estimated differences in the age-adjusted prevalence of self-reported chronic disease and disability and observer-measured health indicators across pseudo-birth cohorts (born <1925, 1925-1935, 1936-1945, 1946-1954, 1955-1959).

**Results:** Age-adjusted prevalence of doctor-diagnosed chronic disease increased across cohorts in all regions. Trends in disability prevalence were more regionally varied. Still, in both the US and Europe, we observed a structural break in disability trends, with declines observed in pre-war cohorts slowing, stalling, or reversing for cohorts born since 1945.

**Discussion:** In all regions, we found evidence for worsening health across cohorts, particularly for those born since 1945. While more chronic disease in younger cohorts need not necessarily translate to worse quality of life or higher rates of functional limitation, there is some suggestion that worsening chronic disease morbidity may be spilling over into worsening disability.

## Introduction

By 2050, nearly one-fifth of the world’s population will be aged ≥65 years due to long-term declines in fertility and increasing life expectancy (LE) (UN, 2020). Population ageing in European countries and the United States (US) has already reached an advanced stage, with 18.6%, 21.1% and 16.8% of the populations of England, the European Union, and the US aged ≥65 years by 2020/21 (Caplan & Rabe, 2023; Eurostat, 2023; ONS, 2023). Population ageing is expected to accelerate as the large Baby Boom generation enters old age. The growing number and proportion of older people in ageing populations have profound societal implications, increasing demand for health and social care (Scott, 2021). These negative effects of population ageing could be mitigated by continued improvements in health and functioning across generations. A key question is whether more recent generations are in better health than previous generations at the same age.

Increases in longevity have long been tied to improvements in population health. Declines in infectious disease mortality largely drove early increases in LE through improved sanitation, nutrition, public health (McKeown & Record, 1962), and medical innovation. Since the 1980s, the prevalence of disability and functional limitations at older ages has declined in many high-income countries (Parker & Thorslund, 2007), suggesting that health and physical functioning were improving across successive generations. While the duration or percentage of life spent with disability and limitations increased in some contexts, this was often due to gains in LE outpacing gains in disability-free LE (Spiers et al., 2021). However, a growing body of research, particularly from the US, suggests that more recently born generations, particularly those born since 1945, may be experiencing worse health than previous generations as measured by chronic disease and disability (Freedman et al., 2013; Martin et al., 2010; Seeman et al., 2010). We refer to this trend of worsening health across cohorts at the same age as the “generational health drift.”

There is some evidence that other high-income countries may also be experiencing a generational health drift for outcomes including disability (e.g., France: Cambois et al., 2013), self-rated health (e.g, England: Jivraj et al., 2020), and chronic disease (e.g., Sweden: Gondek et al., 2021; England: Jivraj et al., 2020; The Netherlands: Oostrom et al., 2016; Spain: Walter et al., 2016). However, combining such findings to understand how generational trends in Europe directly compare to those found in the US is challenging due to the wide variety of health outcomes studied, methods, data sources, and outcome definitions used, and age-groups and periods included in the analyses (Lafortune & Balestat, 2007). For instance, many studies explore trends in the duration of life spent in poor health, which is related to but distinct from whether more recent generations are healthier at the same age and has different implications for public health. Time spent in poor health can increase despite improvements in health at the same age, provided increases in survival are greater than postponement of disease or disability onset. While European countries share many qualities with the US (Western, Educated, Industrialised, Rich, and Democratic), there are also important differences and distinct social, economic, political, and epidemiological histories. Knowing whether and how generational trends in health differ across regions is an important first step to identifying potential drivers of generational health drift, informing strategies to halt or reverse this process.

Research comparing generational trends in health across regions or countries in the same study, which can overcome some of these challenges, has focused on a limited set of outcomes: disability, a single (and severe) form of poor physical health (Chatterji et al., 2015; Fors et al., 2022; Lee et al., 2020; Welsh et al., 2021), and cardiometabolic diseases (Martinson et al., 2022). Some research does suggest that trends in disability might be diverging at younger and older ages, consistent with the idea of generational drift, but trends in chronic disease and disability have not been explored together in the same study. In addition, little research has simultaneously compared generational trends in self-reported and observed disease and disability across regions. Consideration of a broader set of physical health outcomes capturing different aspects of population health can contribute to a better understanding of the factors driving the generational health drift and the societal implications of these trends (e.g., for the labour market, and demand for different areas of health and social care, thereby informing funding allocation across these services (Parker & Thorslund, 2007)).

In this study, we explore generational differences in physical health across birth cohorts born before and after the Second World War in five high-income regions: the US, England, and Western, Northern, and Southern Europe (WE, NE, and SE). Using harmonised data from the Gateway to Global Aging Data, we explore trends for a variety of physical health outcomes, including self-reported doctor-diagnosed chronic disease, disability, and other health indicators. We directly compare these generational trends to explore whether English and European populations have experienced a similar pattern of “generational health drift” as the US across multiple measures of health.

## Methods

### Study Participants

We used data on adults aged ≥51 years from the US, and ≥50 years from England and 11 continental European countries (Austria, Belgium, Denmark, France, Germany, Greece, Italy, The Netherlands, Spain, Sweden, and Switzerland) who participated in the Health and Retirement Study (HRS), the English Longitudinal Study of Ageing (ELSA), or the Survey of Health, Ageing, and Retirement in Europe (SHARE) between 2004 and 2018. HRS, ELSA, and SHARE have similar target populations, frequency and timing of interviews (every two years, except SHARE in 2008 when no health-related outcomes were collected), multidisciplinary questionnaires, and use of refreshment samples to maintain sample representativeness (Sonnega et al., 2014; Steptoe et al., 2013; Börsch-Supan et al., 2013). We used harmonised datasets produced by the Gateway to Global Ageing Data: Harmonised HRS Version C, Harmonised ELSA Version G2 and Harmonised SHARE Version F (Chen et al., 2022; Wilkens et al., 2021, 2022).

The analytical sample was comprised of repeated observations of respondents from HRS, ELSA, and SHARE between 2004 and 2018 who were age-eligible and had a known year of birth. Proxy responses were included where available.

HRS received ethical approval from the University of Michigan Health Sciences/Behavioural Sciences Institutional Review Board, ELSA from the South Central – Berkshire Research Ethics Committee, and SHARE from the Ethics Council of the Max Planck Society and the University of Mannheim. All participants gave informed consent. No further ethics approval was required for this work.

### Exposure

We grouped respondents into five pseudo-birth cohorts, defined by birthyear, roughly corresponding to the Greatest Generation (born before 1925), the early Silent Generation (born 1925-1935), the late Silent Generation (1936-1945), early Baby Boomers (1946-1955), and late Baby Boomers (1955-1959) (Supplementary Figure 1). We used these groupings as we wanted to explore whether generational trends differed for those born before or after the Second World War. Each pseudo-birth cohort consisted of multiple observations of respondents within each birthyear group at different ages.

### Outcomes

We explored trends for self-reported and observer-measured outcomes capturing diverse aspects of physical health: self-reported doctor-diagnosed disease, disability and physical functioning, and other health indicators (self-reported and measured). Additional information on how these outcomes were measured can be found in Supplementary Table 1.

Participants were asked if they had ever been told by a doctor that they had cancer (excluding minor skin cancers), heart problems, chronic lung disease (e.g., chronic bronchitis or emphysema), high blood pressure (BP), or high cholesterol (for ELSA and SHARE). Each outcome was a binary indicator (yes/no) of whether a participant reported having each condition at each wave they participated in.

Participants were also asked about difficulties with six activities of daily living (ADL) related to personal care activities (e.g., eating, bathing), four instrumental activities of daily living (IADL) related to daily tasks (e.g., preparing a hot meal, shopping for groceries), and their ability to perform seven mobility and motor coordination tasks (e.g., walking one block, lifting 10 lbs, picking up a small coin from a flat surface). Using answers to these questions, we constructed four binary indicators to identify respondents with isolated mobility limitations only (≥1 mobility difficulty but no IADL or ADL difficulties), mild disability (≥1 IADL difficulty and any number mobility difficulties but no ADL difficulties), moderate disability (1-2 ADL difficulties and any number of IADL or mobility difficulties), or severe disability (≥3 ADL difficulties and any number of IADL or mobility difficulties) at each wave. These categories were constructed to reflect the idea that mobility limitations do not inevitably result in the inability to perform specific activities, but that there is a high correlation between mobility limitation, IADL limitation, and ADL limitation: a person with ADL limitations is likely to also have IADL and mobility limitations.

We also explored generational trends in continuous maximum grip strength (in kilograms), and binary indicators of obesity (self-reported BMI ≥30 kg/m^2^ for HRS and SHARE and measured BMI ≥30 kg/m^2^ for HRS and ELSA) and measured hypertension (mean systolic BP ≥140 mmHg or mean diastolic BP ≥90 mmHg for HRS and ELSA). While not direct measurements of disease, these are indicators of physical health status correlated with disease and disability risk, and act as useful markers of potential disease or disability risk which can be captured earlier in life.

### Covariates

Covariates in all models were age (year of interview minus birthyear) and gender (man or woman). In models for grip strength, we additionally adjusted for height (in metres) and weight (in kilograms) since grip strength is associated with body size. Calendar year was not included as a covariate since our intent was not to isolate a ‘cohort effect’ but rather to describe differences in disease and disability prevalence accounting for age between generations regardless of whether these resulted from period or cohort effects.

### Data Analysis

We used a pseudo-cohort approach, treating each wave as a cross-sectional sample. We tested the association between birth cohort and each outcome of interest using modified Poisson regression with robust standard errors for binary outcomes (adjusting for age, age^2^, and gender), which has the advantage of producing risk ratios that are more intuitive to interpret than odds ratios (Zou, 2004), and avoids issues of non-collapsibility of the odds ratio (Pang et al., 2016). We explored trends for grip strength using linear regression (adjusting for age, age^2^, gender, height, and weight). Risk ratios were estimated for each country separately using the 1936-1945 cohort as reference group, since this group had the most overlap in age with other pseudo-cohorts, and was one of the largest pseudo-cohorts. We accounted for survey design and clustering of observations within individuals in calculating the standard errors, and used cross-sectional weights and inverse-probability weights for non-response (more details are given below and in the Supplementary Material). We present trends in five regions: the US, England, WE (Austria, Belgium, France, Germany, The Netherlands, and Switzerland), NE (Sweden and Denmark), and SE (Spain, Italy, and Greece). Estimates for countries in continental European regions were statistically pooled using random effects meta-analysis. Data were analysed using Stata (Version 18; StataCorp LP, College Station, TX).

### Missing Data Strategy

Owing to their longitudinal design, HRS, ELSA, and SHARE are affected by attrition which varies by sociodemographic and health status. Even when the data are used cross-sectionally, as we do in this study, results may be biased as the sample’s composition at each cross-section is affected by attrition in the longitudinal sample. We derived inverse-probability weights for non-response at each wave to mitigate the impact of differential non-response. These weights were multiplied with person-level cross-sectional weights provided by the studies, which restore representativeness by gender and age/cohort, to obtain a final analysis weight. We describe the derivation of these weights in the Supplementary Material.

### Sensitivity analyses

We examined results stratified by gender. We also explored how results differed when using cross-sectional weights only, and when additionally adjusting for the total number of waves participants responded to. Using biomarker data from ELSA, we explored generational differences in glycated haemoglobin (HbA1c, a marker of clinical diabetes), total cholesterol, and BP levels accounting for medication. Details are provided in the Supplementary Material.

## Results

The analytical sample was composed of 435,069 observations of 114,526 participants in HRS, ELSA, and SHARE who participated in at least one survey wave between 2004 and 2018 and were age-eligible at the time of interview (Table 1). Birthyear ranged from 1896 to 1959, with a mean birthyear of 1943 (SD = 10 years). The mean number of waves was 3.8, with members of the 1936-1945 cohort participating in the most waves on average (Supplementary Table 2).

**Table 1.** Sociodemographic and health characteristics of the analytical sample (n = 114,526) by birth cohort. Percentages, means, and standard deviations are unweighted. Values correspond to the number of individuals who reported each outcome at least once between 2004 and 2018. Mobility limitation = difficulty with ≥1 mobility task but no IADL or ADL limitation. Mild disability = difficulty with ≥1 IADL but no ADL limitation. Moderate disability = difficulty with 1 or 2 ADL. Severe disability = difficulty with ≥3 ADL. Some individuals had no instances where the outcome was recorded. A description of item non-response is given in Supplementary Tables 2, 3 and 4.

|  | Birth Cohort |  |  |  |  |  |
| --- | --- | --- | --- | --- | --- | --- |
|  | < 1925<br>N (%/mean, SD) | 1925-35<br>N (%/mean, SD) | 1936-45<br>N (%/mean, SD) | 1946-54<br>N (%/mean, SD) | 1955-59<br>N (%/mean, SD) | Total<br>N (%/mean, SD) |
| **Total** | 8423 (100) | 22,223 (100) | 31,547 (100) | 35,922 (100) | 16,411 (100) | 114,526 (100) |
| **Gender** |  |  |  |  |  |  |
| Man | 3150 (37) | 10,275 (46) | 14,963 (47) | 16,886 (47) | 7117 (43) | 52,391 (46) |
| Woman | 5273 (63) | 11,948 (54) | 16,584 (53) | 19,036 (53) | 9294 (57) | 62,135 (54) |
| **Region** |  |  |  |  |  |  |
| US | 3345 (40) | 5792 (26) | 6747 (21) | 6731 (19) | 4324 (26) | 26,939 (23) |
| England | 1220 (14) | 2921 (13) | 4111 (13) | 5022 (14) | 1718 (11) | 14,992 (13) |
| Western Europe | 2022 (24) | 7123 (32) | 11,198 (35) | 13,589 (38) | 5928 (36) | 39,860 (35) |
| Northern Europe | 667 (8) | 2020 (9) | 3435 (11) | 3774 (10) | 1411 (9) | 11,317 (10) |
| Southern Europe | 1159 (14) | 4367 (20) | 6056 (20) | 6806 (19) | 3030 (18) | 21,418 (19) |
| **Education** |  |  |  |  |  |  |
| Degree | 851 (10) | 2991 (14) | 5917 (19) | 8855 (25) | 4363 (27) | 22,977 (20) |
| No degree | 7457 (90) | 18,931 (86) | 25,278 (81) | 26,677 (75) | 11,938 (73) | 90,281 (80) |
| **Doctor diagnoses** |  |  |  |  |  |  |
| Cancer | 1472 (18) | 4214 (19) | 5502 (17) | 4095 (11) | 1344 (8) | 16,627 (15) |
| Heart problems | 3664 (44) | 8449 (38) | 8574 (27) | 5847 (16) | 1840 (11) | 28,374 (25) |
| Lung disease | 1149 (14) | 3423 (15) | 4174 (13) | 3313 (9) | 1317 (8) | 13,376 (12) |
| Diabetes | 1539 (18) | 5308 (24) | 6930 (22) | 6108 (17) | 2388 (15) | 22,273 (19) |
| High blood pressure | 5038 (60) | 14,248 (64) | 18,848 (60) | 17,264 (48) | 6609 (40) | 62,007 (54) |
| High cholesterol | 1404 (25) | 8342 (43) | 13,499 (46) | 13,894 (40) | 5560 (35) | 42,699 (40) |
| **Disability** |  |  |  |  |  |  |
| Mobility limitation | 3963 (47) | 14,745 (66) | 20,326 (65) | 19,870 (56) | 8013 (50) | 66,917 (59) |
| Mild disability | 1932 (23) | 4024 (18) | 2964 (9) | 2242 (6) | 1008 (6) | 12,170 (11) |
| Moderate disability | 3659 (43) | 7625 (35) | 7325 (23) | 5778 (16) | 2292 (14) | 26,829 (23) |
| Severe disability | 2955 (35) | 4321 (19) | 2774 (9) | 2045 (6) | 815 (5) | 12,910 (11) |
| **Biomarkers** |  |  |  |  |  |  |
| Obesity (measured)<sup>1</sup> | 533 (23) | 2677 (40) | 4015 (45) | 4618 (49) | 2455 (51) | 14,298 (44) |
| Obesity (self-reported)<sup>2</sup> | 1054 (15) | 5044 (27) | 8348 (31) | 9466 (31) | 4667 (32) | 28,579 (29) |
| Hypertension<sup>1</sup> | 1064 (43) | 3699 (54) | 5020 (55) | 4675 (48) | 1988 (40) | 24,125 (73) |
| Maximum grip strength (kg)<sup>3</sup> | 5733<br>(23.0, 8.9) | 19,232<br>(29.1, 10.4) | 28,840<br>(34.4, 11.3) | 33,109<br>(37.9, 12.0) | 14,919<br>(38.5, 12.1) | 101,833<br>(34.7, 12.2) |

<sup>1</sup>Only measured in HRS (US) and ELSA (England)<sup>2</sup>Only measured in HRS (US) and SHARE (continental Europe).<sup>3</sup>Mean of the maximum grip strength recorded for each participant across all waves.
*Note:* Percentages, means, and standard deviations are unweighted. Values correspond to the number of individuals who reported each outcome at least once between 2004 and 2018. Mobility limitation = difficulty with $\geq 1$ mobility task but no IADL or ADL limitation. Mild disability = difficulty with $\geq 1$ IADL but no ADL limitation. Moderate disability = difficulty with 1 or 2 ADL. Severe disability = difficulty with $\geq 3$ ADL. Some individuals had no instances where the outcome was recorded. A description of item non-response is given in Supplementary Tables 2, 3 and 4.

### Main Results

#### Self-reported doctor-diagnosed chronic diseases

Overall, the age- and gender-adjusted prevalence of doctor-diagnosed chronic disease increased across successive cohorts in the US and Europe (Figure 1). The prevalence of diabetes and high cholesterol increased in all five regions. Generational trends in cancer, high BP, lung disease, and heart problems also suggested that more recently born cohorts had worse health at the same age. However, there was more regional variation in the magnitude of risk ratios (RRs) and the specific cohorts with the largest RRs. For instance, RRs were systematically larger in SE compared to the US for all six health outcomes, suggesting that prevalence had increased more across cohorts in SE than the US. For cancer, lung disease, and heart problems, which are outcomes capturing many specific conditions, 95% confidence intervals were wider than for specific diagnoses such as diabetes. Still, the overall direction of generational trends was consistent across regions.

**Figure 1.**
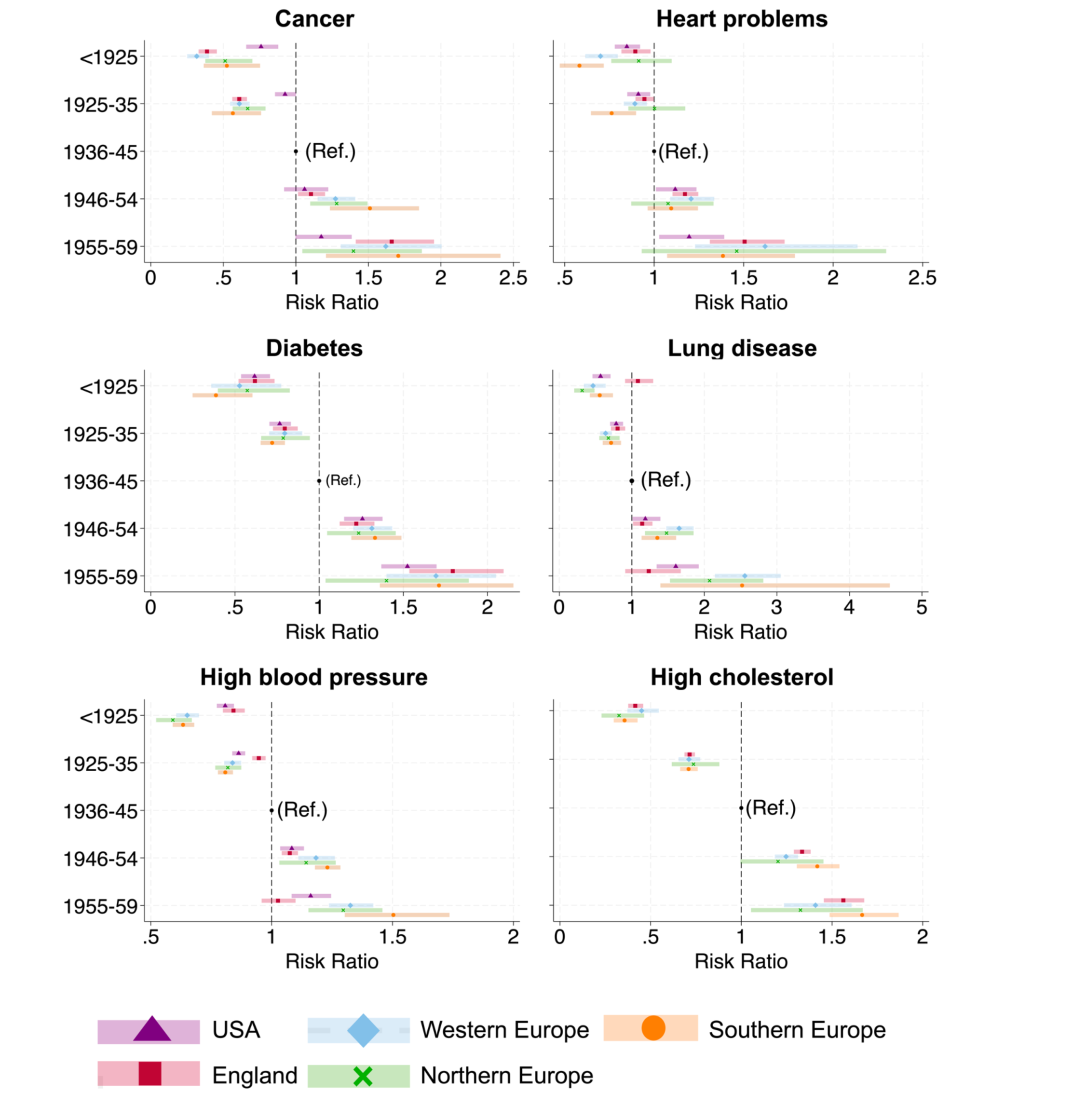
Generational differences in self-reported doctor-diagnosed chronic disease in the United States and Europe. Reference group is the 1936-1945 birth cohort. All results are adjusted for age, age^2^, and gender. Results for the US are not shown for high cholesterol, as information on this outcome was only collected in three sweeps of HRS as opposed to 7 in ELSA and SHARE (Supplementary Table 1). A Risk Ratio (RR) >1 indicates that a cohort has a greater prevalence of the outcome than the reference 1936-1945 cohort accounting for age and gender, while a RR <1 indicates that the cohort had a lower prevalence.

#### Disability and mobility limitation

Trends in disability and mobility limitation were more regionally varied (Figure 2); however, in the US and most European regions, declines in disability prevalence seen in pre-war cohorts appeared to slow, stall, or reverse in post-war cohorts. In the US, the age-adjusted prevalence of isolated mobility limitations (without IADLs or ADLs) declined across cohorts, beginning with the 1925-1936 cohort. There was also evidence for declines in the prevalence of mild disability (IADL without ADLs) and moderate disability (1-2 ADLs) across pre-war cohorts, but the prevalence remained stable or increased again in cohorts born after 1945. The prevalence of severe disability (≥3 ADLs) increased in post-war cohorts. Trends in England were similar, though the prevalence of mild disability remained stable across post-war cohorts rather than reversed and declines in prevalence of moderate disability slowed but did not stall entirely. The shape and direction of generational trends in SE were very similar to those in England, except for severe disability, for which we found no strong evidence of a generational trend. Despite some improvements across pre-war cohorts, there was no strong evidence for further declines in age-adjusted prevalence of mobility limitations or disability across post-war cohorts in WE or NE.

**Figure 2.**
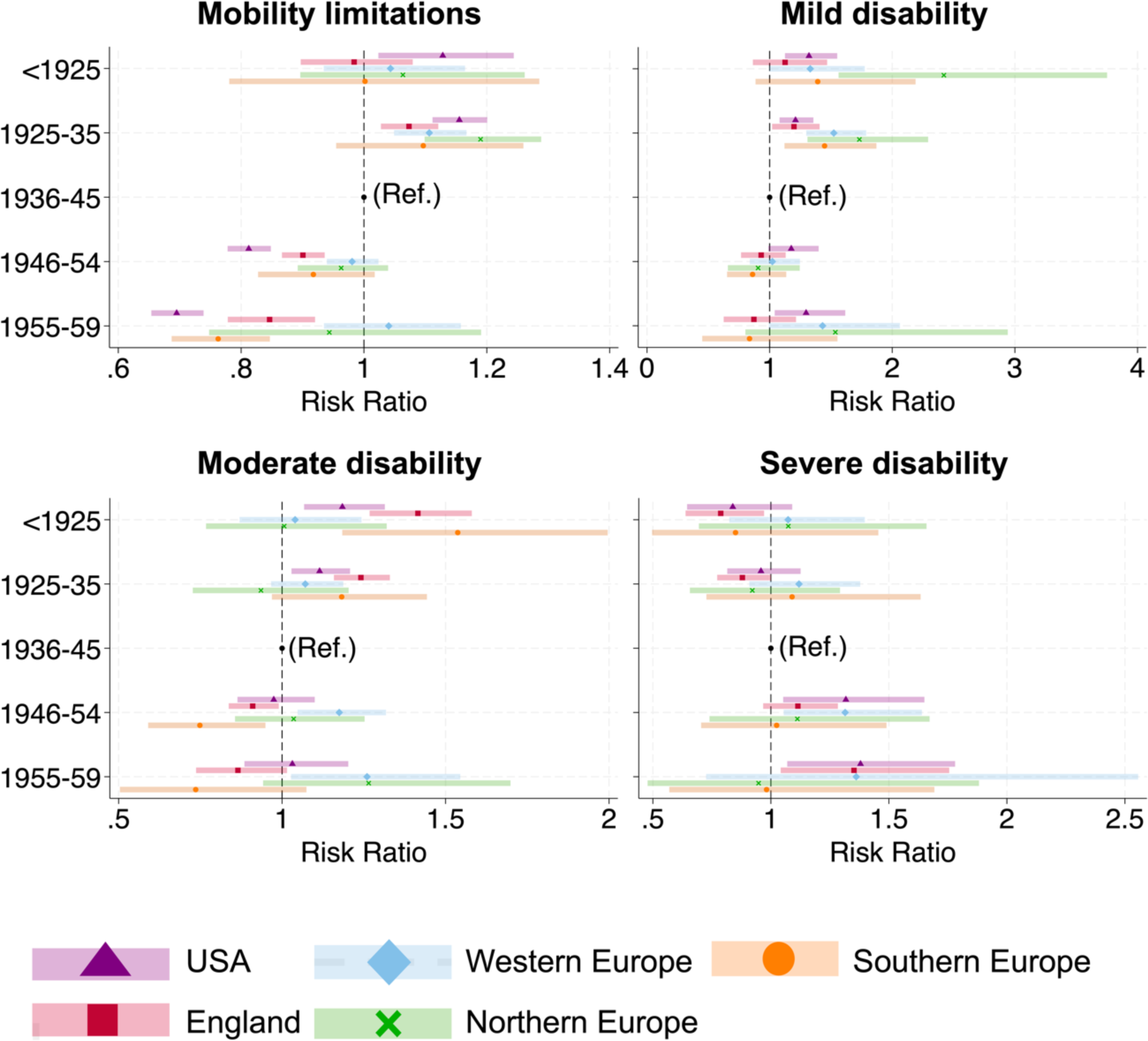
Generational differences in self-reported disability and mobility limitation in the United States and Europe. Reference group is the 1936-1945 birth cohort. All results are adjusted for age, age^2^, and gender. Mobility limitations = respondent reports ≥1 mobility limitation, but no IADL or ADL limitations. Mild disability = respondent reports ≥1 IADL limitation and any number of mobility limitations but no ADL limitations. Moderate disability = respondent reports 1-2 ADL limitations, and any number of IADL or mobility limitations. Severe disability = respondent reports ≥3 ADL limitations, and any number of IADL or mobility limitations. A Risk Ratio (RR) >1 indicates that a cohort has a greater prevalence of the outcome than the reference 1936-1945 cohort accounting for age and gender, while a RR <1 indicates that the cohort had a lower prevalence.

#### Other indicators of physical health

Age-adjusted obesity prevalence increased across cohorts, based on self-reported BMI for the US and continental European regions and measured BMI for England and the US (Figure 3). However, in SE such increases in prevalence were only observed for cohorts born before 1945, and prevalence did not increase further in post-war cohorts. The risk of measured hypertension decreased across cohorts in both the US and England. Trends in age, height, and weight-adjusted maximum grip strength exhibited a high degree of regional variability, with evidence for declines in the US and England, and increases in NE across the full set of cohorts. Estimates for SE and WE showed no clear pattern, though there was some suggestion of declining grip strength in the most recently born cohorts in both regions.

**Figure 3.**
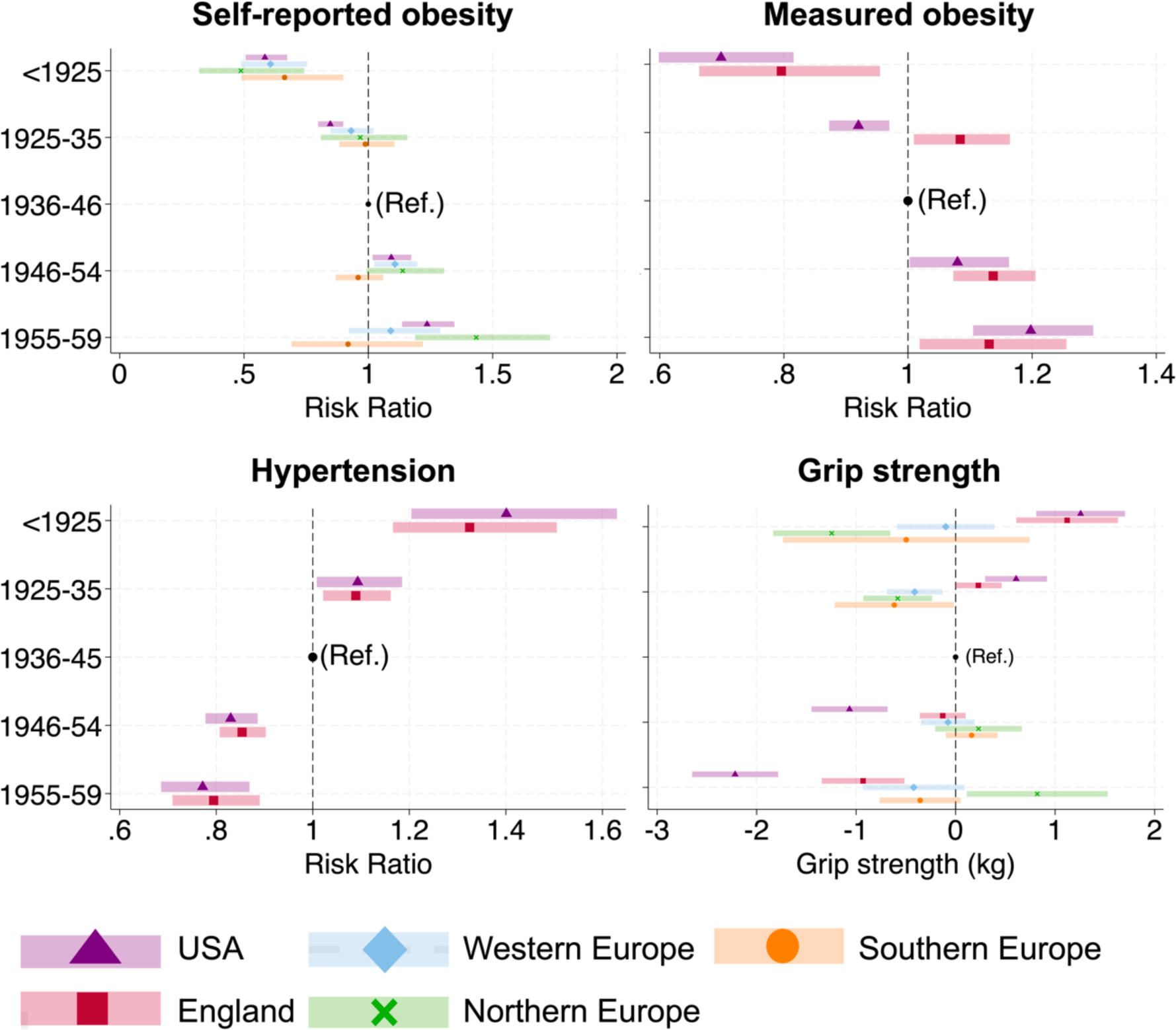
Generational differences in obesity, hypertension, and grip strength in the United States and Europe. Reference group is the 1936-1945 birth cohort. Results for obesity and hypertension are adjusted for age, age^2^ and gender. Results for grip strength are adjusted for age, age^2^, gender, height (in metres) and mass (in kilograms). Results for hypertension and measured obesity are presented for the US and England only. Results for self-reported obesity are shown for the US and continental European regions only. Hypertension = average systolic blood pressure ≥140 mmHg or average diastolic blood pressure ≥90 mmHg. Obesity = measured or self-reported BMI ≥30 kg/m^2^. Grip strength (kilograms) is a continuous outcome. A Risk Ratio (RR) >1 indicates that a cohort has a greater prevalence of the outcome than the reference 1936-1945 cohort accounting for age and gender, while a RR <1 indicates that the cohort had a lower prevalence. For grip strength, a point estimate >0 is indicative of stronger grip strength in a cohort compared to the reference 1936-1945 reference cohort, while a point estimate <0 is indicative of weaker grip strength accounting for age, gender, height, and weight.

### Sensitivity Analyses

Results were similar when stratifying by gender and did not differ significantly when cross-sectional weights only were used or when number of waves participated in were adjusted for (results available upon request). In ELSA, accounting for medication, more recent generations had a higher age-adjusted prevalence of high HbA1c, high total cholesterol, and hypertension (Supplementary Figure 2).

## Discussion

We compared generational trends in physical health and disability for cohorts born before and after the Second World War in the US and Europe to explore whether a similar “generational health drift” was experienced across regions. More recently born cohorts in all regions had a higher prevalence of doctor-diagnosed chronic disease than their predecessors at the same age. Trends in functional limitation were more nuanced and regionally variable. While the age-adjusted prevalence of isolated mobility limitations declined across cohorts in most regions, declines in disability prevalence seen in pre-war cohorts appeared to slow, stall, or reverse for cohorts born after 1945 in the US and most European regions. These findings support the idea that a generational drift in physical health has occurred in both the US and Europe, whereby younger generations have worse health than previous generations at the same age, particularly for cohorts born since 1945.

Our finding that the prevalence of doctor-diagnosed disease has increased across successive cohorts aligns well with findings from other studies. Studies on older adults in the US have found evidence for the increasing prevalence of multimorbidity (Bishop et al., 2022), as well as of specific conditions such as cancer, diabetes, hypertension, lung disease, and heart disease (Beltrán-Sánchez et al., 2016; Payne, 2022), both within age-groups over time and across cohorts. Similarly, the age-standardised period prevalence of many major non-communicable diseases (e.g., diabetes, lung cancer, stroke) in the UK has been increasing since 1946 (Gondek et al., 2019), and a higher risk of chronic disease morbidity has been noted across English post-war cohorts (Jivraj et al., 2020). Worsening chronic disease morbidity across cohorts has also been described in continental European countries (e.g., Spain: Walter et al., 2016; Sweden: Gondek et al., 2021; The Netherlands: Oostrom et al., 2016). We noted that RRs in the US were often smaller than for other regions, which may be explained by changes in dietary, occupational, and smoking patterns beginning earlier in the US than in other regions, resulting in less pronounced changes in exposure to disease risk factors across these cohorts in the US. For instance, male lung cancer mortality, attributable to smoking, began to decline for post-1920s cohorts in the US (Preston & Wang, 2006), but only for post-1950s cohorts in southern Spain (Córdoba-Doña et al., 2023).

The regional variability in generational trends in disability and mobility limitations noted in our study has also been reported for period trends in disability prevalence (Lee et al., 2020). While the prevalence of isolated mobility limitations declined in most regions, trends in disability prevalence in several regions appeared to show a structural break around 1945, declining across pre-war cohorts, but stalling or reversing in post-war cohorts. Examining trends in the prevalence of limitations affecting major activity and work in Americans aged 30-39 years born 1915-1959 (similar to the birthyears included in this study), Reynolds et al. (1998) found that while cohorts born before 1945 progressively became less disabled, prevalence increased again for cohorts born after 1945, suggesting that the generational trends observed in our study among adults aged ≥50 years may already have been apparent in earlier life. Diverging trends in disability at younger and older ages in the US have also been reported between 1988/94-1999/2004 and 2000-2008 (Freedman et al., 2013; Seeman et al., 2010). In England, an absence of declines in disability in post-war cohorts of working-age adults or diverging trends in disability at younger and older ages consistent with our results have also been noted Welsh et al., 2021). Comparing our results to other studies on trends in disability in continental Europe is challenging given the variety of time-periods and age-groups covered in the literature. While studies on older adults (aged ≥65 years) often document declines in IADL, ADL or mobility limitations up to the 2000s (e.g., Norway: Moe & Hagen, 2011; Spain: Zunzunegui et al., 2006), a more recent study capturing post-war cohorts who have entered older age is consistent with our findings of stalling improvements or worsening disability and functional limitation across post-war cohorts (Fors et al., 2022). The generational drift in disability could be explained by many factors, including changing associations between disability and chronic disease morbidity, multimorbidity, and obesity (Chang et al., 2017; Sperlich et al., 2021). Contextual differences in the treatment and management of these chronic conditions and in educational and occupational changes across cohorts may explain regional differences in disability trends.

Trends in other health indicators can help to contextualise trends in self-reported outcomes. The increase in age-adjusted prevalence of obesity, a risk factor for cardiometabolic disease, is consistent with increasing prevalence of outcomes such as diabetes across cohorts. We found that age-adjusted prevalence of obesity remained stable for those born after 1936 in SE, which could be explained by the compensatory effect of rapidly increasing stature across these cohorts (Garcia & Quintana-Domeque, 2007). Improvements in nutrition driving changes in childhood growth patterns and adult stature could also explain continued increases in grip strength across post-war cohorts (Kuh et al., 2006), and the lack of worsening disability in SE. Trends in grip strength by region were broadly consistent with findings from single-country studies from countries within those regions. For instance, grip strength improved across cohorts of Finnish adults aged ≥75 years born between the 1910s and 1940s, consistent with our findings for NE (Koivunen et al., 2021). In age-period-cohort models, Beller et al. (2019) found evidence that grip strength remained stable across birth cohorts in Sweden accounting for age and period, while Germans born 1930-1945 had stronger grip strength than previous and subsequent generations, Spaniards experience increasing grip strength across cohorts born up to the 1960s, at which point grip strength declined. These patterns are consistent with our findings since we did not adjust for period. Such regional differences likely reflect differences in the balance of nutritional improvements and declines in physical activity (Dodds et al., 2013), related to changes in occupation. Since lower grip strength in midlife is associated with higher risk of disability and mobility limitation in later life (Soysal et al., 2021), declining grip strength across US and English birth cohorts suggests that younger generations may reach older ages in a frailer state and at higher risk of functional impairment.

To understand the possible implications of these generational trends in health, it is important to know whether worse self-reported health at the same age in younger generations reflects truly worse underlying health. Trends in self-reported outcomes can result from several mechanisms including changes in medical and diagnosis practices, changes in health-awareness and reporting style across generations (e.g., due to decreasing stigma in reporting certain health conditions), and true worsening health. While untangling the contribution of these factors is complex, triangulating evidence from self-reported and measured outcomes using a consistent methodological approach, as we have done in this study, can give insight into whether the generational drift in health can be explained away by phenomena other than worsening underlying health.

We illustrate this using the example of high BP in England. The prevalence of self-reported doctor-diagnosed high BP increased across successive cohorts, although increases decelerated across post-war cohorts, possibly because of opposing trends in obesity and smoking. Policies incentivising BP monitoring in primary care (e.g., Quality and Outcomes Framework) and screening programmes (e.g., the National Health Service Health Check) may result in more people with undiagnosed high BP receiving a diagnosis of hypertension, such that individuals with high BP born more recently likely receive diagnoses earlier in life. The prevalence of measured hypertension decreased across cohorts, but after accounting for medication use the generational trends in measured hypertension were consistent with those for doctor-diagnosed high BP. Biomarker measures can be affected by measurement error and selection bias (since not all participants consent to nurse visits) but are not affected by changes in health awareness or demand for diagnosis. Triangulating evidence from three BP outcomes therefore suggests that in England, younger generations have higher BP than previous generations at the same age, but that their BP is better managed, which may lower risk of more lethal and disabling conditions like cardiovascular and cerebrovascular disease.

Supplementary analyses also showed that more recent generations of older adults in England had a greater prevalence of high HbA1c and total cholesterol, consistent with increasing prevalence of doctor-diagnosed diabetes and high cholesterol in more recent generations. Zheng and Echave (2021) have similarly found evidence for greater physiological dysregulation across cohorts of US adults born since 1945. These findings support that, while the impact of changing diagnosis, health awareness, and reporting style cannot be ruled out, generational trends in doctor-diagnosed chronic conditions such as diabetes and high BP do reflect a true generational health drift.

While our measures of disability and mobility rely on self-reports, we do not think it likely that the stalling or reversing declines in disability in most regions can be entirely explained through changing reporting styles. Difficulty performing IADLs and ADLs is a function of both physical capability and the environment (e.g., access to technology). Environmental changes are likely to have led to less difficulty performing tasks, particularly IADLs, such that younger cohorts are likely to underreport difficulty for a similar underlying degree of impairment. An exception to this may be changes in intergenerational cohabitation, with more recent cohorts of older adults having to perform more of these tasks without support resulting in more difficulties being reported. Mobility limitations are measured by asking respondents to report whether they can perform specific physical actions, and it is unlikely that there would be systematic generational differences in how these questions are answered.

Our work does not explicitly relate trends in physical health with trends in mortality. However, since the birth cohorts included in this study are extinct or already relatively aged, we can hypothesise about what trends in age-adjusted prevalence of poor health may mean for length of life in poor health. Provided cohort LE continues to increase or remains stable, worse health in younger generations at the same age will result in in more years spent with poor physical health, driven both by continued increases in survival and by declining age of onset of morbidity, in contradiction with the compression of morbidity hypothesis (Fries, 1980). This interpretation should be made with care, since further increases in cohort LE for the most recent birth cohorts included in our analysis are uncertain. In the US, there is some suggestion that worsening health in younger cohorts is impacting LE. Slowing or stalling declines in cardiometabolic disease-related mortality for cohorts born since the 1950s have contributed to the worsening all-cause mortality rates among middle-aged white Americans alongside increases in drug-related deaths (Masters et al., 2018). If the generational health drift does result in more years spent in poor health, this will have considerable implications for health and social care expenditure and for how funding is allocated across these services. Worsening health in younger cohorts at the same age also has important employment policy implications, as national governments move to raise the State Pension Age to respond to the challenge of population ageing.

Strengths of this study include its use of large-scale harmonised data from multiple countries, of a standardised methodological approach, and that we explicitly addressed the potential impact of selection bias on cross-sectional samples by developing inverse probability weights for non-response. Another strength is the wide range of health outcomes explored, allowing findings to be triangulated for a better understanding the potential societal implications of generational trends in physical health. This study also has limitations. While HRS, ELSA and SHARE follow similar designs, some variability between studies remains (e.g., sampling frame, timing of refreshment samples, not all outcomes measured at all waves). Our use of harmonised data meant that some outcomes (heart problems, lung disease, cancer) were broad and included conditions which may be evolving in different directions, making the interpretation of these trends challenging.

Our results suggest that populations in the US and Europe may be experiencing a generational health drift, with younger generations exhibiting worse health than previous generations at the same age for variety of physical health outcomes, particularly for cohorts born after 1945. This drift was observed in all regions for chronic disease, while evidence for generational drift in disability was more regionally varied. Worsening quality of life and increasing disability and functional limitation are not inevitable consequences of increasing chronic disease in younger generations, provided these conditions are well-managed. Concerningly, our results suggest that previous declines in disability have stalled or reversed in the US and some European regions, suggesting that despite improvements in medical knowledge and technology, worsening chronic disease status in these younger cohorts may be spilling over to disability. More research is needed to better understand the drivers of the generational health drift, which may vary by context. A lifecourse approach could improve our understanding of when in life generational differences in physical and mental health emerge, thereby contributing to the understanding of the mechanisms behind the generational health drift, and crucially, informing strategies to reverse it.

## Acknowledgments

HRS is sponsored by the National Institute on Aging (grant number U01AG009740) and the Social Security Administration. ELSA is funded by the National Institute on Aging (grant number R01AG017644); and by a consortium of UK government departments (the Department for Health and Social Care; the Department for Transport; and the Department for Work and Pensions) which is coordinated by the National Institute for Health Research (grant number 198-1074). Funding has also been provided by the Economic and Social Research Council (ESRC). The SHARE project received funding from the European Commission Directorate General for Research and Innovation; the Directorate General of Social Affairs and Inclusion; and additional funding from the German Ministry of Education and Research; the Max Planck Society for the Advancement of Science; the National Institute on Aging (U01_AG09740-13S2, P01_AG005842, P01_AG08291, P30_AG12815, R21_AG025169, Y1-AG-4553-01, IAG_BSR06-11, OGHA_04-064, HHSN271201300071C, RAG052527A); and various other national funding sources.

## Author contributions

LG conceptualised the study, together with GBP, AG, and JBD. LG conducted the statistical analyses. GBP supervised the statistical analyses. All authors contributed to the interpretation of the results. LG wrote the first draft of the manuscript. GBP, AG and JBD critically revised the manuscript.

## Data Availability

All data used in this study are publicly accessible to researchers. Harmonised datasets for HRS, ELSA, and SHARE, and accompanying codebooks can be accessed via the Gateway to Global Aging Data website (www.g2gad.org). The additional HRS data used in this study can be accessed through the HRS website (www.hrsdata.isr.umich.edu).

## Ethical Approval and Consent to Participate

HRS received approval from the University of Michigan Health Sciences/Behavioural Sciences Institutional Review Board. ELSA received ethical approval from the South Central – Berkshire Research Ethics Committee. SHARE received ethical approval from the Ethics Committee of the Max Planck Society for the Advancement of Science at the University of Mannheim. All participants gave informed consent at recruitment.

