## Supplementary for "Generational differences in physical health and disability in the United States and Europe"

### Supplementary Material

**Tables**


1 Description of variables used to construct outcome measures. 2
2 Distribution of observations by age group and survey wave. 3
3 Description of item non-response by survey wave for HRS 4
4 Description of item non-response by survey wave for ELSA 5
5 Description of item non-response by survey wave for SHARE 6
6 Distribution of observations by cohort and survey wave 7
7 Imputed predictors of wave non-response 11


**Figures**


1 Lexis diagram showing pseudo-cohorts 8
2 Generational differences in blood pressure, blood cholesterol and HbA1c in ELSA 9


**Supplementary Text**


1 Inverse probability weights for non-response 10
2 Accounting for complex survey design 12

**Supplementary Table 1.** *Description of variables used to construct outcome measures.*

| Outcome | Waves collected | | | Description | Response |
|---|---|---|---|---|---|
| | HRS | ELSA | SHARE | | |
| **Functional limitations** | | | | | |
| ADL limitation | All | All | All except 2008 | Difficulty with dressing, bathing, eating, getting in/out of bed, using the toilet, walking across a room. | Yes/No |
| IADL limitation | All | All | All except 2008 | Difficulty with preparing a hot meal, shopping for groceries, taking medication, managing money. | Yes/No |
| Mobility limitation | All | All | All except 2008 | Difficulty with walking one block, climbing several flights of stairs without rest, getting up from a chair, stooping/kneeling/crouching, reaching overhead, lifting 10 lbs, picking up a small coin from a table. | Yes/No |
| **Doctor diagnoses** | | | | | |
| Cancer | All | All | All except 2008 | Ever told by a doctor they had cancer or a malignant tumour, excluding minor skin cancers and including blood disorders such as lymphoma and leukaemia. | Yes/No |
| Heart problems | All | All | All except 2008 | Ever told by a doctor they had heart problems. This includes conditions like heart attacks, coronary heart disease, angina, congestive heart failure, heart murmurs, and coronary thrombosis. | Yes/No |
| Lung disease | All | All | All except 2008 | Ever told by a doctor they had chronic lung disease such as chronic bronchitis or emphysema. | Yes/No |
| High blood pressure | All | All | All except 2008 | Ever told by a doctor they had high blood pressure or hypertension. | Yes/No |
| High cholesterol | - | All | All except 2008 | Ever told by a doctor that their blood cholesterol level was high. | Yes/No |
| Diabetes | All | All | All except 2008 | Ever told by a doctor they had diabetes or high blood sugar. | Yes/No |
| **Biomarkers** | | | | | |
| Obesity (measured)[1] | All | - | All except 2008 | BMI $\geq$ 30 kg/m$^2$ based on observer-measured height and weight. | Yes/No |
| Obesity (self-reported)[1] | All | 2004, 2008, 2012, 2016 | - | BMI $\geq$ 30 kg/m$^2$ based on self-reported height and weight. | Yes/No |
| Body mass index | All | 2004, 2008, 2012, 2016 | All except 2008 | We included body mass index in models for grip strength. Self-reported BMI was used for HRS and SHARE, while measured BMI was used for ELSA. | Continuous |
| Grip strength | All | All | All except 2008 | Maximum hand grip strength in dominant hand across two (HRS, SHARE) or three (ELSA) readings, using handheld dynamometer. | Continuous |
| Systolic BP[2] | All except 2004 | 2004, 2008, 2012, 2016 | - | Average systolic blood pressure over three readings using an automated blood pressure monitor. | Continuous |
| Diastolic BP[2] | All except 2004 | 2004, 2008, 2012, 2016 | - | Average diastolic blood pressure over three readings using an automated blood pressure monitor. | Continuous |
| **ELSA Supplementary Analyses** | | | | | |
| HbA1c | - | 2004, 2008, 2012, 2016 | - | From blood samples collected during nurse visits. In DCCT (%) units for 2004 and 2008, and in IFCC (mmol/mol) units converted to DCCT units for 2012 and 2016. | Continuous |
| Total cholesterol | - | 2004, 2008, 2012, 2016 | - | From blood samples collected during nurse visits. In mg/dL. | Continuous |
| Systolic BP[2] | - | 2004, 2008, 2012, 2016 | - | Average systolic blood pressure over three readings using an automated blood pressure monitor. | Continuous |
| Diastolic BP[2] | - | 2004, 2008, 2012, 2016 | - | Average diastolic blood pressure over three readings using an automated blood pressure monitor. | Continuous |
| Antihypertensive drugs | - | 2004, 2008, 2012, 2016 | - | Whether respondent takes medication for high blood pressure, or medication that prevents them from getting high blood pressure anymore (from 2008). Asked if report diagnosis of high blood pressure. | Yes/No |
| Drugs for cholesterol | - | 2008, 2012, 2016 | - | Whether respondent is taking medication for high cholesterol. Asked if reported diagnosis of high cholesterol. | Yes/No |
| Drugs for diabetes | - | 2004, 2008, 2012, 2016 | - | Whether respondent takes oral medication or uses insulin shots for diabetes. Asked if reported a diabetes or high blood sugar diagnosis. | Yes/No |

[1] BMI cut-offs for obesity are based on World Health Organisation (2021) standards.

[2] Systolic blood pressure <70 mmHg or >270 mmHg and diastolic blood pressure <50 mmHg or >150 mmHg were considered implausible and recoded as missing. Based on cut-offs used by (Zhou *et al.* (2017)

**Supplementary Table 2.** *Distribution of observations by cohort and survey wave.*

| | Birth cohort | | | | | |
|---|---|---|---|---|---|---|
| | **< 1925** | **1925-35** | **1936-45** | **1946-54** | **1955-59** | **Total** |
| **2004** | 6900 | 14,622 | 17,736 | 15,963 | 43 | 55,264 |
| **2006** | 5356 | 13,382 | 17,031 | 16,597 | 2512 | 54,878 |
| **2008** | 2591 | 6766 | 8830 | 7943 | 1218 | 24,348 |
| **2010** | 3105 | 12,237 | 18,437 | 20,725 | 10,285 | 64,789 |
| **2012** | 2445 | 12,496 | 20,669 | 24,142 | 12,315 | 72,067 |
| **2014** | 1596 | 10,545 | 18,694 | 22,241 | 11,340 | 64,416 |
| **2016** | 857 | 8035 | 16,216 | 19,872 | 10,181 | 55,161 |
| **2018** | 349 | 5122 | 12,248 | 15,633 | 7,794 | 41,146 |
| ***N* observations** | 23,199 | 83,205 | 129,861 | 143,116 | 55,688 | 435,069 |
| ***N* respondents** | 8423 | 22,223 | 31,547 | 35,922 | 16,441 | 114,526 |
| **Obs/Resp, Mean (SD)** | 2.8 (2.8) | 3.7 (2.3) | 4.1 (2.4) | 4.0 (2.3) | 3.4 (1.6) | 3.8 (2.2) |

*Note:* The number of observations in 2008 is smaller since only HRS and ELSA collected health-related variables in this year. These counts are based on participation in core interviews. The number of respondents is smaller for outcomes based on measurements taken during enhanced face-to-face interviews in HRS or nurse interviews in ELSA (e.g., obesity based on measured height and weight, hypertension based on measured systolic and diastolic blood pressure, grip strength).

**Supplementary Table 3.** *Description of item non-response by survey wave for HRS.*

| | 2004 | 2006 | 2008 | 2010 | 2012 | 2014 | 2016 | 2018 |
|---|---|---|---|---|---|---|---|---|
| *N core interview* | *18,700* | *17,105* | *15,805* | *20,337* | *18,850* | *17,092* | *15,125* | *12,414* |
| *N nurse visit*[1] | *3271* | *7167* | *6421* | *8463* | *7940* | *7432* | *6045* | *5270* |
| | **N (%)** | **N (%)** | **N (%)** | **N (%)** | **N (%)** | **N (%)** | **N (%)** | **N (%)** |
| **Cohort** | 0 | 0 | 0 | 0 | 0 | 0 | 0 | 0 |
| **Gender** | 0 | 0 | 0 | 0 | 0 | 0 | 0 | 0 |
| **Education** | 3 (0.02) | 3 (0.02) | 3 (0.02) | 4 (0.02) | 3 (0.02) | 4 (0.02) | 4 (0.03) | 3 (0.02) |
| **Age** | 0 | 0 | 0 | 0 | 0 | 0 | 0 | 0 |
| **Disability** | | | | | | | | |
| Limitation | 20 (0.11) | 14 (0.08) | 14 (0.09) | 122 (0.60) | 20 (0.11) | 13 (0.08) | 15 (0.10) | 23 (0.19) |
| Mild | 16 (0.09) | 13 (0.08) | 11 (0.07) | 118 (0.58) | 19 (0.10) | 13 (0.08) | 15 (0.10) | 22 (0.18) |
| Moderate | 8 (0.04) | 9 (0.05) | 8 (0.05) | 111 (0.55) | 16 (0.08) | 12 (0.07) | 14 (0.09) | 17 (0.14) |
| Severe | 8 (0.04) | 9 (0.05) | 8 (0.05) | 111 (0.55) | 16 (0.08) | 12 (0.07) | 14 (0.09) | 17 (0.14) |
| **Doctor diagnoses** | | | | | | | | |
| Heart problem | 0 | 0 | 0 | 0 | 0 | 0 | 0 | 0 |
| Lung disease | 0 | 0 | 0 | 0 | 0 | 0 | 0 | 0 |
| Cancer | 0 | 0 | 0 | 0 | 0 | 0 | 0 | 0 |
| Diabetes | 0 | 0 | 0 | 0 | 0 | 0 | 0 | 0 |
| High BP | 0 | 0 | 0 | 0 | 0 | 0 | 0 | 0 |
| **Biomarkers** | | | | | | | | |
| High measured BP | NA | 258 (3.60) | 258 (4.02) | 395 (4.67) | 336 (4.23) | 224 (3.01) | 144 (2.38) | 142 (2.69) |
| Obesity (SR) | 316 (1.69) | 250 (1.46) | 206 (1.30) | 379 (1.86) | 270 (1.43) | 253 (1.48) | 207 (1.37) | 170 (1.37) |
| Obesity (measured)[2] | 2765 (84.53) | 468 (6.53) | 394 (6.14) | 525 (6.20) | 501 (6.31) | 464 (6.24) | 370 (6.12) | 375 (7.12) |
| BMI (SR) | 325 (1.74) | 258 (1.51) | 216 (1.37) | 216 (1.37) | 306 (1.62) | 266 (1.56) | 215 (1.42) | 176 (1.42) |
| Grip strength | 283 (8.65) | 334 (4.66) | 265 (4.13) | 443 (5.23) | 468 (5.89) | 498 (6.70) | 404 (6.68) | 388 (7.36) |
| **Extra Covariates** | | | | | | | | |
| Height (SR) | 10 (0.05) | 6 (0.04) | 5 (0.03) | 48 (0.24) | 40 (0.21) | 40 (0.23) | 32 (0.21) | 33 (0.27) |
| Weight (SR) | 271 (1.45) | 239 (1.40) | 194 (1.23) | 267 (1.31) | 189 (1.00) | 197 (1.15) | 163 (1.08) | 124 (1.00) |

[1] This corresponds to the number of individuals who participated in the physical measurement modules of HRS at each wave. An alternating half of the HRS sample received in-depth enhanced face-to-face interviews at each wave, during which anthropometric and biomarker measures are taken, and physical performance tests are conducted. Weights provided by HRS aim to restore the representativeness of this half-sample to the target population.

[2]Observer-measured height and weight in 2004 were collected for a subset of individuals who were part of the enhanced face-to-face interview, as part of an experimental module, explaining the higher amount of missing data on measured obesity in 2004.

*Note:* This table shows the number and percentage of missing observations by wave and variable, for individuals who participated in each wave of HRS between 2004 and 2018 and had a non-zero analysis weight. This table thus quantifies item non-response (i.e., missing data among those who responded to each survey wave). BMI = Body Mass Index. BP = Blood Pressure. SR = Self-Reported.

**Supplementary Table 4.** *Description of item non-response by survey wave for ELSA.*

| | 2004 | 2006 | 2008 | 2010 | 2012 | 2014 | 2016 | 2018 |
|---|---|---|---|---|---|---|---|---|
| *N core interview* | *8780* | *8655* | *9659* | *8988* | *8748* | *7598* | *6679* | *5892* |
| *N nurse interview* | *7666* | *NA* | *8195* | *NA* | *7445* | *NA* | *5418* | *NA* |
| | **N (%)** | **N (%)** | **N (%)** | **N (%)** | **N (%)** | **N (%)** | **N (%)** | **N (%)** |
| **Cohort** | 0 | 0 | 0 | 0 | 0 | 0 | 0 | 0 |
| **Gender** | 0 | 0 | 0 | 0 | 0 | 0 | 0 | 0 |
| **Education** | 769 (8.76) | 696 (8.04) | 765 (7.92) | 714 (7.94) | 672 (7.68) | 567 (7.46) | 499 (7.47) | 454 (7.71) |
| **Age** | 0 | 0 | 0 | 0 | 0 | 0 | 0 | 0 |
| **Disability** | | | | | | | | |
| Limitation | 3 (0.03) | 5 (0.06) | 4 (0.04) | 7 (0.08) | 4 (0.05) | 2 (0.03) | 2 (0.03) | 4 (0.07) |
| Mild | 2 (0.02) | 3 (0.03) | 4 (0.04) | 6 (0.07) | 3 (0.03) | 2 (0.03) | 0 | 4 (0.07) |
| Moderate | 2 (0.02) | 3 (0.03) | 4 (0.04) | 6 (0.07) | 3 (0.03) | 2 (0.03) | 0 | 4 (0.07) |
| Severe | 2 (0.02) | 3 (0.03) | 4 (0.04) | 6 (0.07) | 3 (0.03) | 2 (0.03) | 0 | 4 (0.07) |
| **Diagnoses** | | | | | | | | |
| Heart problem | 0 | 0 | 0 | 0 | 0 | 0 | 0 | 0 |
| Lung disease | 0 | 0 | 0 | 0 | 0 | 0 | 0 | 0 |
| Cancer | 0 | 0 | 0 | 0 | 0 | 0 | 0 | 0 |
| Diabetes | 0 | 0 | 0 | 0 | 0 | 0 | 0 | 0 |
| Cholesterol | 4 (0.05) | 0 | 0 | 1 (0.01) | 1 (0.01) | 0 | 0 | 0 |
| High BP | 0 | 0 | 0 | 0 | 0 | 0 | 0 | 0 |
| **Biomarkers** | | | | | | | | |
| High measured BP | 143 (1.87) | NA | 146 (1.78) | NA | 112 (1.50) | NA | 76 (1.40) | NA |
| Obesity (measured) | 441 (5.75) | NA | 359 (4.38) | NA | 337 (4.53) | NA | 283 (5.22) | NA |
| BMI (measured) | 441 (5.75) | NA | 360 (4.39) | NA | 337 (4.53) | NA | 283 (5.22) | NA |
| Grip strength | 156 (2.03) | NA | 206 (2.51) | NA | 225 (3.02) | NA | 263 (4.85) | NA |
| **Extra Covariates** | | | | | | | | |
| Height (measured) | 216 (2.82) | NA | 84 (1.03) | NA | 42 (0.56) | NA | 131 (2.42) | NA |
| Weight (measured)[1] | 293 (3.82) | NA | 201 (2.45) | NA | 213 (2.86) | NA | 15 (0.28) | NA |

[1]Weight measured in 2016 enriched with data on weight measured in 2018 if available.

*Note:* This table shows the number and percentage of missing observations by wave and variable, for individuals who participated in each wave of HRS between 2004 and 2018 and had a non-zero analysis weight. This table thus quantifies item non-response (i.e., missing data among those who responded to each survey wave). BMI = Body Mass Index. BP = Blood Pressure. SR = Self-Reported.

**Supplementary Table 5.** *Description of item non-response by survey wave for SHARE.*

| | 2004 | 2006 | 2010 | 2012 | 2014 | 2016 | 2018 |
|---|---|---|---|---|---|---|---|
| *N core interview* | *26,754* | *27,542* | *33,929* | *42,805* | *38,186* | *32,083* | *21,739* |
| *N nurse visit*[1] | *26,754* | *27,542* | *33,929* | *42,805* | *38,186* | *32,083* | *21,739* |
| | **N (%)** | **N (%)** | **N (%)** | **N (%)** | **N (%)** | **N (%)** | **N (%)** |
| **Cohort** | 0 | 0 | 0 | 0 | 0 | 0 | 0 |
| **Gender** | 0 | 0 | 0 | 0 | 0 | 0 | 0 |
| **Education** | 0 | 0 | 0 | 0 | 0 | 0 | 0 |
| **Age** | 0 | 0 | 0 | 0 | 0 | 0 | 0 |
| **Disability** | | | | | | | |
| Limitation | 142 (0.53) | 91 (0.33) | 179 (0.53) | 109 (0.25) | 81 (0.21) | 417 (1.30) | 63 (0.29) |
| Mild | 134 (0.50) | 88 (0.32) | 174 (0.51) | 107 (0.25) | 78 (0.20) | 323 (1.01) | 60 (0.28) |
| Moderate | 134 (0.50) | 88 (0.32) | 174 (0.51) | 107 (0.25) | 78 (0.20) | 323 (1.01) | 60 (0.28) |
| Severe | 134 (0.50) | 88 (0.32) | 174 (0.51) | 107 (0.25) | 78 (0.20) | 323 (1.01) | 60 (0.28) |
| **Diagnoses** | | | | | | | |
| Heart problem | 79 (0.30) | 65 (0.24) | 116 (0.34) | 72 (0.17) | 48 (0.13) | 97 (0.30) | 43 (0.20) |
| Lung disease | 73 (0.27) | 68 (0.25) | 116 (0.34) | 79 (0.18) | 52 (0.14) | 100 (0.31) | 49 (0.23) |
| Cancer | 75 (0.28) | 69 (0.25) | 116 (0.34) | 81 (0.19) | 51 (0.13) | 107 (0.33) | 50 (0.23) |
| Diabetes | 78 (0.29) | 70 (0.25) | 118 (0.35) | 77 (0.18) | 49 (0.13) | 102 (0.32) | 45 (0.21) |
| Cholesterol | 79 (0.30) | 67 (0.24) | 116 (0.34) | 76 (0.18) | 40 (0.10) | 98 (0.31) | 36 (0.17) |
| High BP | 91 (0.34) | 71 (0.26) | 127 (0.37) | 72 (0.17) | 43 (0.11) | 82 (0.26) | 26 (0.12) |
| **Biomarkers** | | | | | | | |
| Obesity (SR) | 570 (2.13) | 810 (2.94) | 1455 (4.29) | 1092 (2.55) | 1055 (2.76) | 906 (2.82) | 472 (2.17) |
| BMI (SR) | 578 (2.16) | 817 (2.97) | 1465 (4.32) | 1112 (2.60) | 1063 (2.78) | 916 (2.86) | 482 (2.22) |
| Grip strength | 2590 (9.68) | 2830 (10.28) | 3472 (10.23) | 4288 (10.02) | 3304 (8.65) | 3353 (10.45) | 1821 (8.38) |
| **Extra Covariates** | | | | | | | |
| Height (SR) | 260 (0.97) | 277 (1.01) | 323 (0.95) | 137 (0.32) | 105 (0.27) | 78 (0.24) | 57 (0.26) |
| Weight (SR) | 393 (1.47) | 388 (1.41) | 657 (1.94) | 696 (1.63) | 674 (1.77) | 850 (2.65) | 423 (1.95) |

[1] Similar to HRS, physical measures in SHARE (physical performance, grip strength) were collected during the core interview visit, and not during a separate interview as is the case in ELSA.

*Note:* This table shows the number and percentage of missing observations by wave and variable, for individuals who participated in each wave of HRS between 2004 and 2018 and had a non-zero analysis weight. This table thus quantifies item non-response (i.e., missing data among those who responded to each survey wave). In 2008, SHARE did not collect any health-related data. BMI = Body Mass Index. BP = Blood Pressure. SR = Self-Reported.

**Supplementary Table 6.** *Distribution of observations by cohort and age-group.*

| | Birth cohort | | | | | |
|---|---|---|---|---|---|---|
| **Age group** | **< 1925** | **1925-35** | **1936-45** | **1946-54** | **1955-59** | **Total** |
| **50-54** | 0 | 0 | 0 | 14,637 | 15,858 | 30,495 |
| **55-59** | 0 | 0 | 1979 | 35,708 | 27,480 | 65,167 |
| **60-64** | 0 | 0 | 17,874 | 48,299 | 12,286 | 78,459 |
| **65-69** | 0 | 2179 | 36,647 | 36,285 | 63 | 75,174 |
| **70-74** | 0 | 15,332 | 41,996 | 8187 | 1 | 65,516 |
| **74-79** | 290 | 26,610 | 26,125 | 0 | 0 | 53,025 |
| **80-84** | 6574 | 25,150 | 5240 | 0 | 0 | 36,964 |
| **85+** | 16,335 | 13,934 | 0 | 0 | 0 | 30,269 |
| ***N* observations** | 23,199 | 83,205 | 129,861 | 143,116 | 55,688 | 435,069 |
| ***N* respondents** | 8423 | 22,223 | 31,547 | 35,922 | 16,441 | 114,526 |

*Note:* These counts are based on participation in core interviews. The number of respondents is smaller for outcomes based on measurements taken during enhanced face-to-face interviews in HRS or nurse interviews in ELSA (e.g., obesity based on measured height and weight, hypertension based on measured systolic and diastolic blood pressure, grip strength).

**Supplementary Figure 1.** *Lexis diagram showing pseudo-cohorts.*

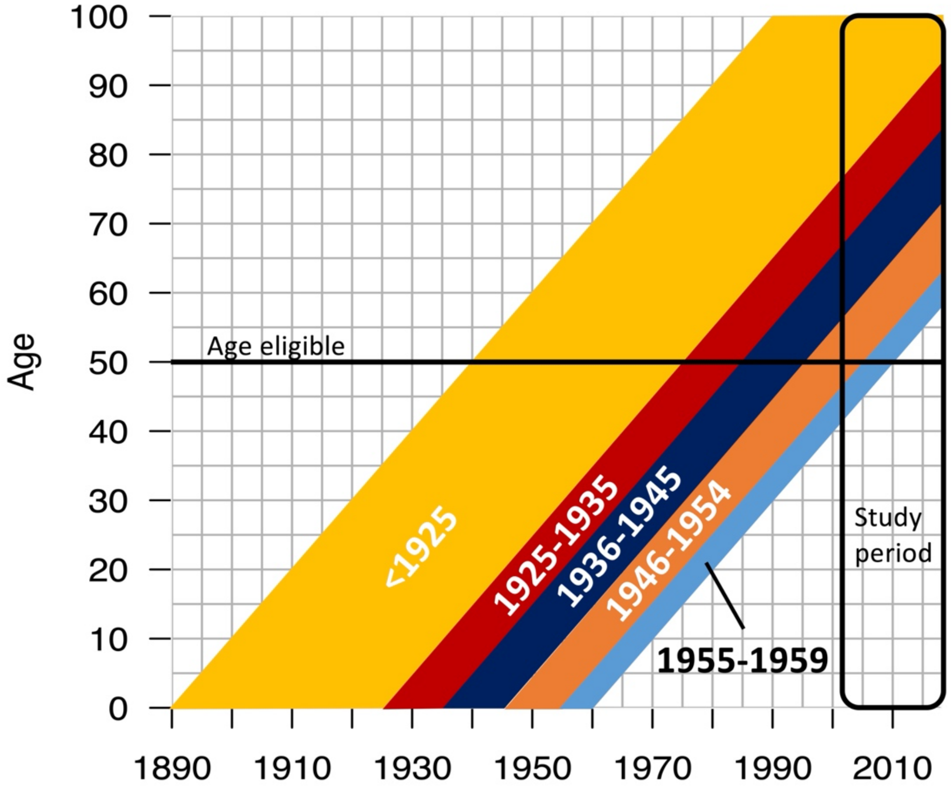

**Supplementary Figure 2.** *Generational differences in blood pressure, blood cholesterol, and HbA1c in ELSA.*

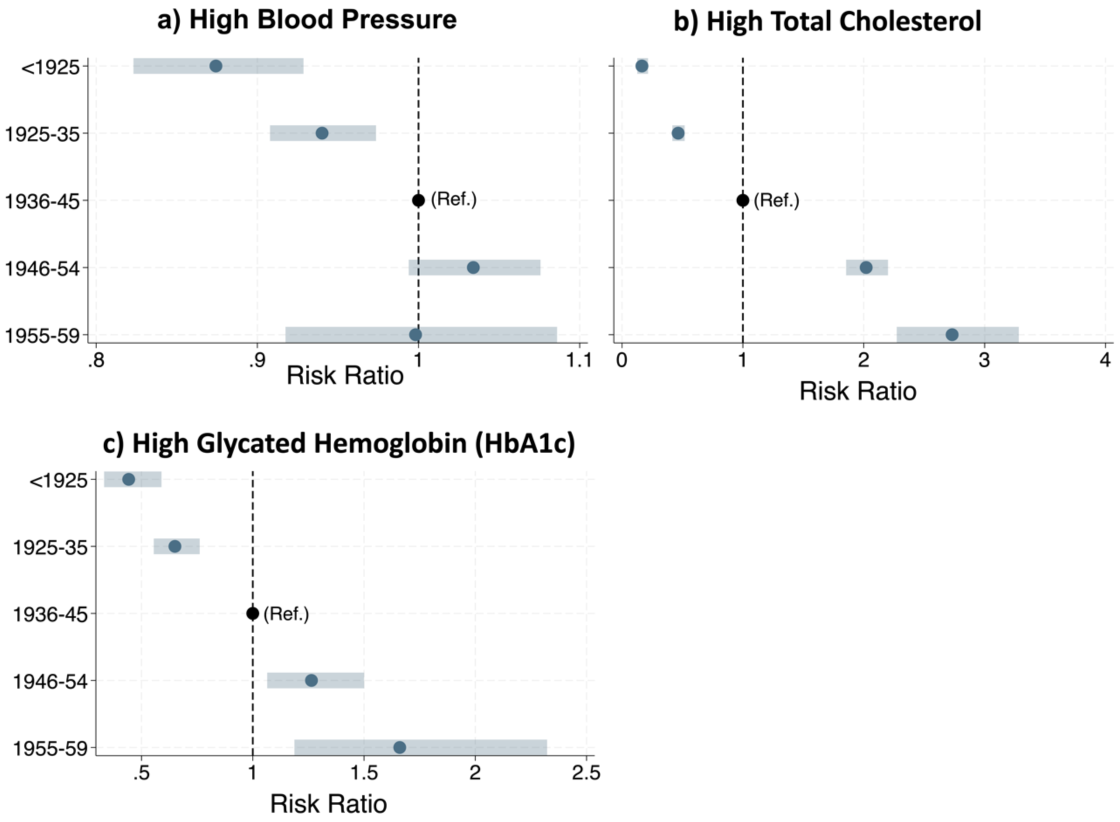


*Note:* All models are unstratified and adjust for age and gender. We used inverse probability weights for non-response multiplied with weights derived for blood samples by the ELSA study for total cholesterol and HbA1c measures, and inverse probability weights multiplied by survey weights for nurse interviews for blood pressure. (a) Respondents were considered to have high blood pressure (BP) if they had systolic BP ≥ 140 mmHg or diastolic BP ≥ 90 mmHg or reported taking drugs to manage their blood pressure. (b) Respondents were considered to have high HbA1c (an indicator of clinical diabetes) if HbA1c ≥ 6.5%. An adjustment of +1 percentage-point for HbA1c was made for respondents who reported taking medication to manage their diabetes (either injected insulin or oral medication). (c) Respondents were considered to have high blood cholesterol if total cholesterol ≥ 200 mg/dL (5.2 mmol/L), or if they reported taking drugs to manage their cholesterol levels.

**Supplementary Text 1.** *Inverse probability weights for unit non-response*

Like all longitudinal studies, SHARE, ELSA, and HRS are affected by attrition, that is, respondents who previously participated in the study stop responding, either because they die or because they drop out of the study. The likelihood of attrition from these studies is not the same for all individuals. It is differential across many characteristics including age, gender, demographic characteristics, socioeconomic position, physical and mental health, and geographic location. Non-response in SHARE, HRS, and ELSA has been shown to be associated with health status. Among younger age-groups better health appears to be associated with a higher rate of non-response, while in older age-groups the relationship is inverse or no association between health and response is observed (Banks et al., 2011; Muszyńska-Spielauer & Spielauer, 2022).

Owing to the longitudinal nature of the data, even when the data are used cross-sectionally as we do in this study, most respondents in each cross-section are part of a longitudinal sample who have responded to prior waves. While the studies do use refreshments to maintain representativeness, they do not occur at every wave, occur at different times and with different frequency across countries, and usually focus on restoring sample size at younger ages. As such, the composition of the sample at each cross-sectional wave is affected by attrition of the longitudinal sample. While HRS, SHARE, and ELSA do provide person-level cross-sectional analysis weights which restore sample representativeness at each wave to the age and sex structure of the target population in that survey year, these weights do not account for differential non-response by factors other than gender, age/cohort, and geography. Since our outcome of interest was physical health status, and health status is known to be associated with non-response, it is important for us to account for differential attrition by health status in our analyses.

Data missingness in longitudinal studies like SHARE, ELSA and HRS is primarily driven by wave non-response and attrition rather than unit non-response (i.e., not answering a particular question or set of questions in an interview). Because of this, we chose to derive inverse probability weights for non-response at each wave which could be multiplied by the cross-sectional analysis weights provided alongside the survey data.

We identified a priori variables which are known to be associated with survey non-response. These included sociodemographic variables (educational attainment, gender, age, marital status and whether respondents had children) and health-related variables which were asked at every survey wave and for all studies: doctor-diagnosed conditions (diabetes, cancer, high blood pressure, heart problems, and lung disease), indictors of functioning (any IADL limitation, any ADL limitation and any mobility difficulties), and self-rated general health (Supplementary Table 7).

We generated 25 imputed datasets using the multiple imputation by chained equations. The long form dataset contained observations for individuals who were aged ≥ 50 at the time of the wave and excluded observations for SHARE in 2008 since no health-related data was collected in that year. The imputation model included the outcome (whether participant responded to wave), incomplete predictors of non-response shown in Supplementary Table 7, an indicator of person ID, country, region, study, age, $age^2$, cohort, birthyear, the cross-sectional weights provided by the studies which carry information on strata and clustering, the total number of core and nurse waves that respondents had participated in, and an interaction term between country and cohort. The model imputed variables for men and women separately. All variables with missing data were binary and were imputed using logit.

**Supplementary Table 7.** *Imputed predictors of wave non-response.*

| Variable | Type | Definition |
|---|---|---|
| Education | Binary | Degree (ISCED-97 levels 5 and 6); no degree (ISCED-97 levels 0 to 4) |
| Marital status | Binary | Currently married or cohabitating; divorced/separated/never married. |
| Childbearing | Binary | Whether has any living children. |
| IADL limitation | Binary | Whether experiences difficulty with ≥ 1 IADL activity.[1] |
| ADL limitation | Binary | Whether experiences difficulty with ≥ 1 ADL activity.[1] |
| Mobility limitation | Binary | Whether experiences difficulty with ≥ 1 mobility task.[1] |
| Diabetes | Binary | Ever diagnosed with diabetes/high blood sugar. |
| Cancer | Binary | Ever diagnosed with cancer (excluding minor skin cancers). |
| High BP | Binary | Ever diagnosed with high blood pressure/hypertension. |
| Heart problems | Binary | Ever diagnosed with heart problems.[1] |
| Lung disease | Binary | Ever diagnosed with chronic lung disease. |
| Self-rated health | Binary | Excellent/very good/good; fair/poor. |

[1] See supplementary Table 1 for more detail on the variable definitions.

We imputed data for 816,711 observations, of which 50% had some missing data. After imputing missing values, we reshaped the data into wide format, and built logistic regression models with indicators of non-response to the core interview at each wave from 2006 onwards as the outcome. We constructed separate models for each study since SHARE did not collect any health-related data in 2008. The models contain the variables in the table above, as well as country (for SHARE), gender, age, birthyear, and total number of waves. Predictors of non-response are variables collected at previous waves, including whether an individual participated in a prior wave. Using these models, we predicted the probability of responding to each wave among those who did participate based on their other characteristics. We generate weights as the reciprocal of the probability of response at each wave. We explored different truncation values by exploring how standard estimates varied depending on the truncation value. We truncated weights at 50 (54 weights truncated).

The cross-sectional analysis weight provided by the study was retained in its original form for each study participant's first response to the study from 2004 onward. All participants who participated in SHARE. ELSA or HRS in 2004 therefore has an IPW = 1 such that the final weight was the original analysis weight provided by the study. A participant who first entered the study in 2010 as part of a refreshment sample would also have IPW = 1. For all subsequent waves that a participant responded to, the weight for that observation was obtained by multiplying the cross-sectional analysis weights provided by the study with IPW. When a participant did not participate in the study, the weight for that observation is 0.

The multiply imputed data was only used to derive inverse-probability weights. The analysis featured in this study used the original, unimputed datasets, in combination with these weights.

**Supplementary Text 2.** *Accounting for complex survey design.*

To account for survey design in the analyses, ELSA, SHARE and HRS all provide variables on primary sampling units (PSU) and primary strata.

Sampling design in SHARE differs between countries depending on the sampling frame available. In Denmark for example, respondents were sampled from the national population register using a simple random sampling approach. As such Danish respondents in the harmonised SHARE dataset do not have any information on PSU or strata. Some other countries, such as France, only had data on PSU and primary strata for the core sample and some refreshment samples, but not others. To be able to analyse the data in a coordinated way, we populated missing strata using a constant value within countries within countries, and a unique value of PSU equal to participant ID. This most closely emulates a simple random sampling design, with no stratification and no clustering.

Italy, Spain, and Switzerland had information on both primary and secondary strata and sampling units. We explored whether accounting for this second level of stratification/clustering in addition to the primary strata and clusters and the person ID identifier impacted our results. We found only very small differences in the size of the standard errors. To maintain consistency across analytical models, we limited survey design variables to primary strata, PSUs, and an indicator for within-individual clustering. We add an indicator of person ID as a second level of clustering since we are treating observations from different cross-sections as independent of one another (i.e., no longitudinal modelling), but want to reflect the fact that observations come from individuals measured over time in the calculation of the standard errors.

#### References from Supplementary Material

Banks, J., Muriel, A., & Smith, J. P. (2011). Attrition and health in ageing studies: Evidence from ELSA and HRS. *Longitudinal and Life Course Studies*, *2*(2), 10.14301/llcs.v2i2.115.

Muszyńska-Spielauer, M., & Spielauer, M. (2022). Cross-sectional estimates of population health from the survey of health and retirement in Europe (SHARE) are biased due to health-related sample attrition. *SSM - Population Health*, *20*, 101290. h

World Health Organisation. (2021, June 9). *Obesity and overweight*. World Health Organisation. https://www.who.int/news-room/fact-sheets/detail/obesity-and-overweight

Zhou, B., Bentham, J., Cesare, M. D., Bixby, H., Danaei, G., Cowan, M. J., Paciorek, C. J., Singh, G., Hajifathalian, K., Bennett, J. E., Taddei, C., Bilano, V., Carrillo-Larco, R. M., Djalalinia, S., Khatibzadeh, S., Lugero, C., Peykari, N., Zhang, W. Z., Lu, Y., … Cisneros, J. Z. (2017). Worldwide trends in blood pressure from 1975 to 2015: A pooled analysis of 1479 population-based measurement studies with 19·1 million participants. *The Lancet*, *389*(10064), 37–55.
